# Algorithms for the identification of prevalent diabetes in the All of Us Research Program validated using polygenic scores – a new resource for diabetes precision medicine

**DOI:** 10.1101/2023.09.05.23295061

**Authors:** Lukasz Szczerbinski, Ravi Mandla, Philip Schroeder, Bianca C. Porneala, Josephine H. Li, Jose C. Florez, Josep M. Mercader, Alisa K. Manning, Miriam S. Udler

## Abstract

**OBJECTIVE:** The study aimed to develop and validate algorithms for identifying people with type 1 and type 2 diabetes in the All of Us Research Program (AoU) cohort, using electronic health record (EHR) and survey data.

**RESEARCH DESIGN AND METHODS:** Two sets of algorithms were developed, one using only EHR data (EHR), and the other using a combination of EHR and survey data (EHR+). Their performance was evaluated by testing their association with polygenic scores for both type 1 and type 2 diabetes.

**RESULTS:** For type 1 diabetes, the EHR-only algorithm showed a stronger association with T1D polygenic score (*p*=3×10^−5^) than the EHR+. For type 2 diabetes, the EHR+ algorithm outperformed both the EHR-only and the existing AoU definition, identifying additional cases (25.79% and 22.57% more, respectively) and showing stronger association with T2D polygenic score (DeLong *p*=0.03 and 1×10^−4^, respectively).

**CONCLUSIONS:** We provide new validated definitions of type 1 and type 2 diabetes in AoU, and make them available for researchers. These algorithms, by ensuring consistent diabetes definitions, pave the way for high-quality diabetes research and future clinical discoveries.

**GRAPHICAL ABSTRACT:** 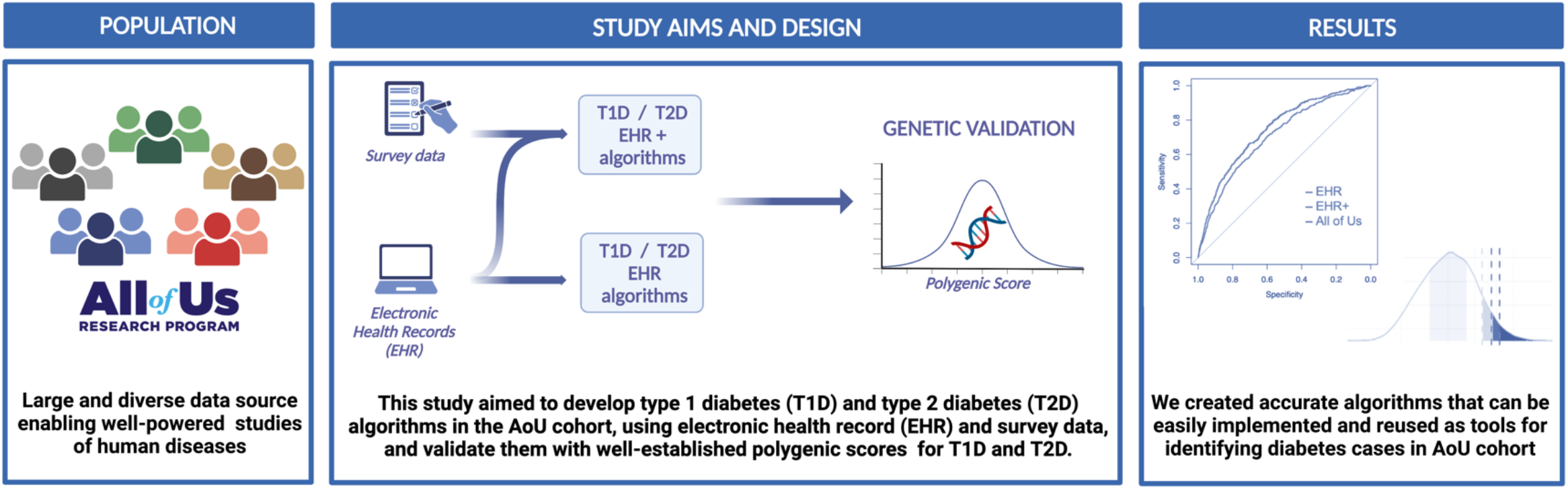

**Article Highlights:** *Why did we undertake this study?:* This study was conducted to develop and validate algorithms for identifying type 1 and type 2 diabetes cases in the All of Us Research Program (AoU).

*What is the specific question(s) we wanted to answer?:* Can accurate algorithms for type 1 and type 2 diabetes identification be developed and validated using AoU cohort Electronic Health Record (EHR) and survey data? Do the identified diabetes cases show association with polygenic scores in diverse populations?

*What did we find?:* We developed a new validated type 1 diabetes definition and expanded upon the existing type 2 diabetes definition.

*What are the implications of our findings?:* The developed algorithms can be universally implemented in AoU for identifying study participants for well-defined case-control diabetes studies.

## 1. Introduction

The All of Us Research Program (AoU) aims to collect data from at least one million individuals across the United States, creating a diverse health database for epidemiological and genomic studies (1). However, the lack of a readily available type 1 diabetes algorithm and the underutilization of all data sources in the existing type 2 diabetes algorithm limit the potential for AoU to contribute to diabetes research. This study addresses these gaps by developing and validating the first type 1 diabetes algorithm and an optimized type 2 diabetes algorithm in AoU, using both electronic health record and survey data. We validated these algorithms using polygenic scores (PSs) (2), assessing their performance across diverse ancestries within the AoU cohort. This work enhances the utility of AoU for high-quality diabetes research and future clinical discoveries.

### 2. Materials and methods

We analyzed data from 372,397 AoU participants enrolled by January 1, 2022, with EHR data available for 309,974 participants and whole genome sequencing (WGS) data for 98,590 participants (extracted from the AoU v6 dataset on November 3, 2022). Genetic ancestry classifications were defined by the AoU Research Program, resulting in subgroups labeled as African/African American (AFR), American Admixed/Latino (AMR), East Asian (EAS); European (EUR) and South Asian (SAS). For the development of diabetes algorithms, we incorporated multiple data points available in AoU, including EHR-based diagnosis, diabetes medications, laboratory data, and self-reported diabetes diagnosis from survey data. More details on the cohort description and data extraction and processing can be found in the **Supplementary Methods**.

We developed algorithms to identify individuals diagnosed with type 1 and 2 diabetes, along with individuals without a diabetes diagnosis, for use as “cases” and “controls” in diabetes studies. Two algorithm versions were created: EHR and EHR+. The EHR algorithms utilized EHR-driven diagnosis, medications, and laboratory measurement values and were developed based on previously reported algorithms (3,4). Modifications were made to ensure relevance to American Diabetes Association’s diagnostic criteria (5) and to exclude certain patient categories to avoid misclassification. The EHR+ algorithms also included self-reported diagnosis obtained from survey data. For the type 1 diabetes case identification algorithm, we modified the algorithm proposed by the eMERGE Phase-IV Program (3) (**Figure 1A**). For the type 2 diabetes case identification algorithm, we modified the Northwestern University algorithm (4) (**Figure 1B**). Moreover, we applied the algorithm for type 2 diabetes case identification available in the “Phenotype Library” of the AoU Researcher Workbench “Featured Workspaces” (labeled ‘AoU-T2D’), to compare the performance of our developed algorithms. Finally, we developed a universal algorithm for the identification of control individuals without diabetes (**Figure 1C**), based on the Northwestern University type 2 diabetes control algorithm (4). Detailed descriptions of the algorithms used in our study are provided in the **Supplementary Methods**.

**Figure 1.**
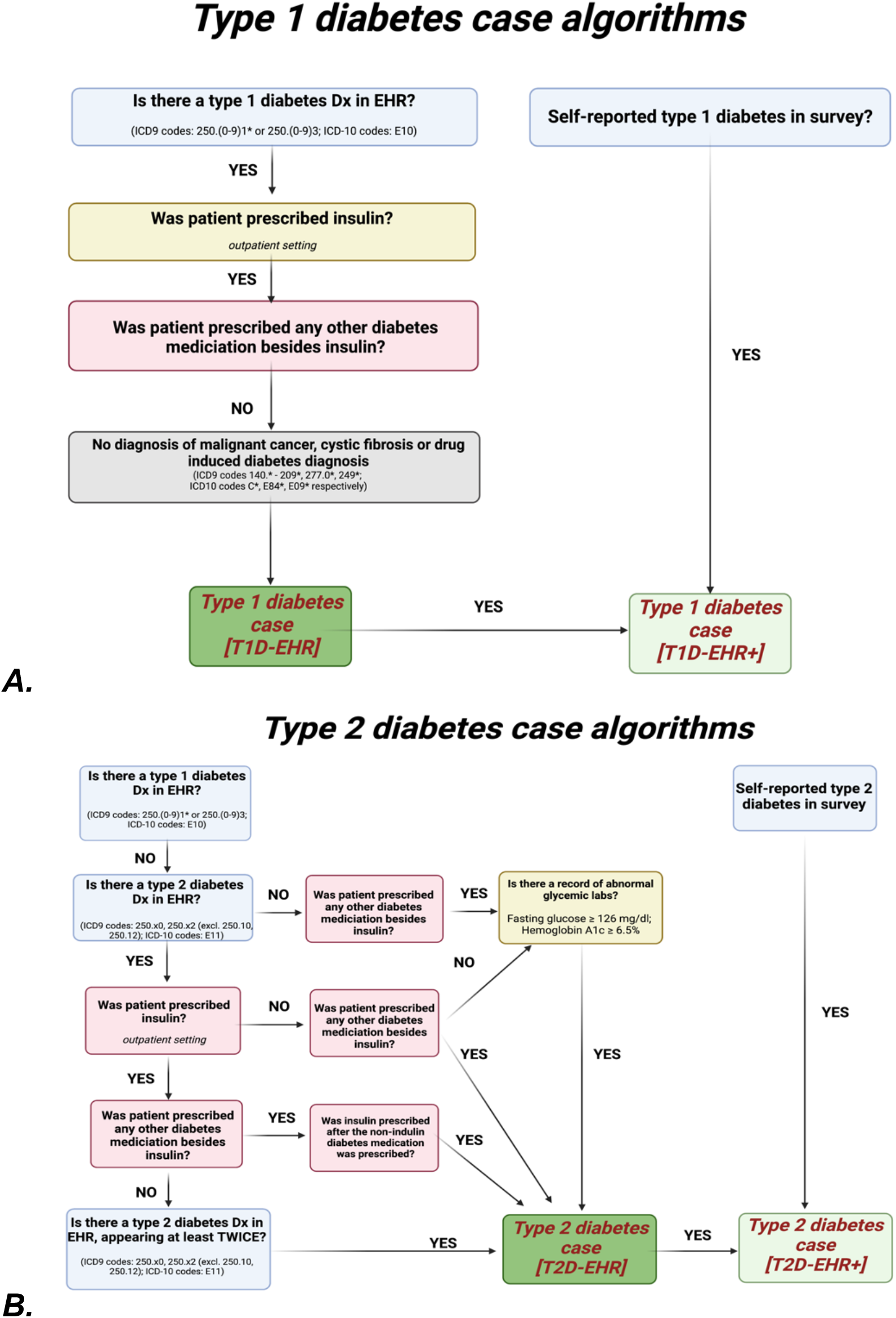
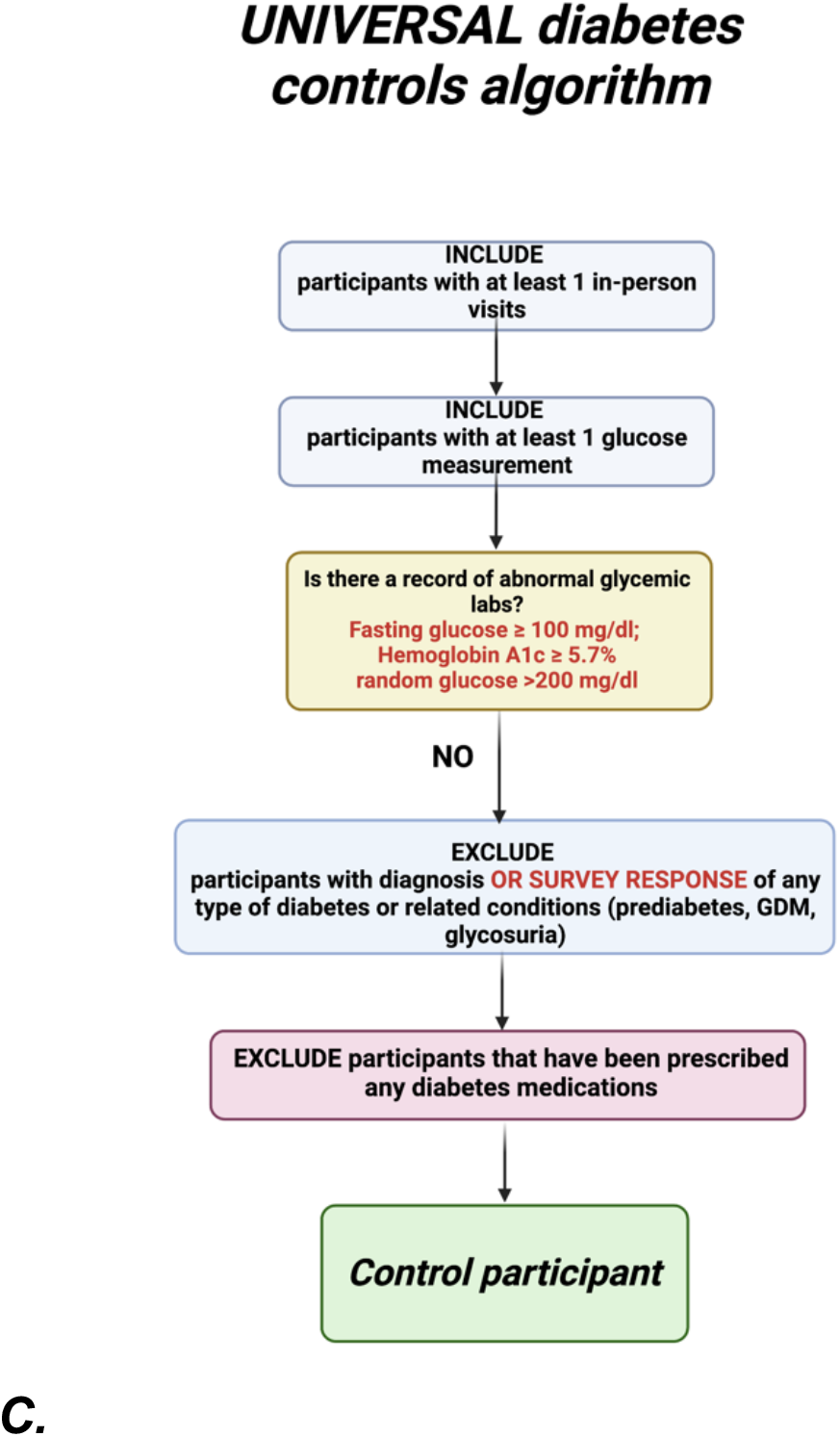
Algorithms for the identification of: A. type 1 diabetes cases; B. type 2 diabetes cases; C. controls without diabetes.

We implemented two published type 1 diabetes polygenic scores: ‘T1D-PS EUR’ from Sharp *et al*. (6), which consisted of 67 single nucleotide polymorphisms (SNPs), derived from European ancestry cohorts; and ‘T1D-PS AA’ from Onengut-Gumuscu *et al*. (7) with 12 variants derived from genetic associations in African-American cohorts. We also created a global extended type 2 diabetes score ‘T2D-PS EUR’, using PRS-CS software, based on a meta-analysis of summary statistics from the European ancestry cohorts (8,9). All scores underwent ancestry adjustment as per methods described by Khera *et al*. (10), using AoU genetic ancestry probabilities. We performed logistic regression analyses to evaluate the accuracy of our diabetes definitions using disease-specific polygenic scores. The analysis included an assessment of the area under the receiver operating characteristic curve (AUC), incremental AUC, which is the difference between the AUC of the full model including the PS and the model only including the covariates, and an evaluation using the DeLong test (11) to compare AUC curves. We also studied the impact of the polygenic scores in the top 10%, 5%, and 2.5% of the distribution compared to the interquartile range. Further details on calculations and statistical analyses are in the **Supplementary Methods**.

## 3. Results

Demographics of the individuals with diabetes identified using the T2D-EHR, T2D-EHR+, T1D-EHR and T1D-EHR+ algorithms are summarized in **Table 1**. Corresponding information for cases identified by the existing AoU-T2D definition and for controls without diabetes identified by our algorithm are presented in **Supplementary Tables S2 and S3**, respectively. To determine the best-performing algorithm for type 1 diabetes and type 2 diabetes, we calculated the associations between diabetes case-control definitions and relevant polygenic scores, and compared odds ratios (ORs), AUCs and incremental AUCs (**Table 2, Figures 2A and 2B**).

**Table 1.** Demographics of diabetes status in AoU, from the diabetes phenotyping algorithms, stratified by self-reported race.

| Type 1 Diabetes |  |  |  |  |  |  |  |  |  |  |  |  |
| --- | --- | --- | --- | --- | --- | --- | --- | --- | --- | --- | --- | --- |
|  | T1D-EHR |  |  |  |  |  | T1D-EHR+ |  |  |  |  |  |
|  | All | NHB | HIS | ASIAN | NHW | Other | All | NHB | HIS | ASIAN | NHW | Other |
| **Number** | 835 | 100 | 129 | N <20 | 538 | 50 | 2457 | 483 | 322 | 43 | 1431 | 178 |
| **Age, mean (SD)** | 49.66 (16.7) | 51.02 (17.0) | 42.25 (15.0) | NA | 51.27 (16.6) | 52.38 (16.0) | 53.73 (16.3) | 54.47 (17.0) | 47.83 (15.5) | 44.14 (13.6) | 54.97 (16.0) | 54.8 (16.6) |
| **BMI kg/m<sup>2</sup>, mean (SD)** | 28.06 (6.9) | 26.73 (5.2) | 27.67 (7.1) | NA | 28.55 (7.3) | 27.23 (5.2) | 30.28 (8.1) | 29.53 (7.4) | 30.39 (8.0) | 25.94 (5.3) | 30.54 (8.3) | 31.04 (8.8) |
| **Female, N (%)** | 470 (56.3%) | 55 (55.0%) | 78 (60.5%) | NA | 309 (57.4%) | 16 (32.0%) | 1382 (56.3%) | 257 (53.2%) | 209 (65.0%) | 26 (60.5%) | 835 (58.4%) | 55 (30.9%) |
| **HbA1c %, mean (SD) [mmol/mol, mean (SD)]** | 7.92 (2.11) [63.11 (23.04)] | 7.62 (1.89) [59.74 (20.68)] | 7.73 (1.95) [60.95 (21.32)] | NA | 8.19 (2.27) [66.03 (24.78)] | 7.55 (1.68) [58.99 (18.34)] | 7.72 (2.03) [60.82 (22.19)] | 7.44 (1.77) [57.78 (19.31)] | 7.72 (1.96) [60.91 (21.47)] | 7.21 (1.18) [55.27 (12.92)] | 7.9 (2.21) [62.82 (24.14)] | 7.41 (1.67) [57.48 (18.2)] |
| Type 2 Diabetes |  |  |  |  |  |  |  |  |  |  |  |  |
|  | T2D-EHR |  |  |  |  |  | T2D-EHR+ |  |  |  |  |  |
|  | All | NHB | HIS | ASIAN | NHW | Other | All | NHB | HIS | ASIAN | NHW | Other |
| **Number** | 28753 | 4059 | 6292 | 603 | 15966 | 1833 | 36172 | 5824 | 7220 | 774 | 19843 | 2511 |
| **Age, mean (SD)** | 63.05 (12.6) | 65.55 (12.5) | 58.94 (12.6) | 62.61 (13.3) | 64.04 (12.2) | 63.06 (13.1) | 63.11 (12.6) | 65.64 (12.3) | 58.75 (12.6) | 62.27 (13.2) | 63.99 (12.1) | 63.09 (13.2) |
| **BMI kg/m<sup>2</sup>, mean (SD)** | 34.06 (8.2) | 34.11 (8.2) | 33.21 (7.7) | 29.35 (6.8) | 34.62 (8.4) | 33.43 (8.0) | 34.09 (8.2) | 34.06 (8.1) | 33.32 (7.6) | 29.44 (7.0) | 34.62 (8.3) | 33.62 (8.0) |
| **Female, N (%)** | 16235 (56.5%) | 1949 (48.0%) | 3952 (62.8%) | 317 (52.6%) | 9267 (58.0%) | 750 (40.9%) | 20187 (55.9%) | 2855 (49.0%) | 4547 (63.0%) | 397 (51.3%) | 11462 (57.8%) | 926 (36.9%) |
| **HbA1c %, mean (SD) [mmol/mol, mean (SD)]** | 7.35 (1.9) [56.8 (20.82)] | 7.13 (1.62) [54.41 (17.75)] | 7.28 (1.85) [56.1 (20.24)] | 7.47 (1.91) [58.19 (20.86)] | 7.43 (1.99) [57.76 (21.79)] | 7.25 (1.86) [55.73 (20.29)] | 7.34 (1.9) [56.75 (20.72)] | 7.12 (1.62) [54.37 (17.72)] | 7.27 (1.83) [55.96 (20.01)] | 7.43 (1.88) [57.76 (20.6)] | 7.43 (1.99) [57.75 (21.73)] | 7.25 (1.85) [55.74 (20.22)] |

**Table 2.**
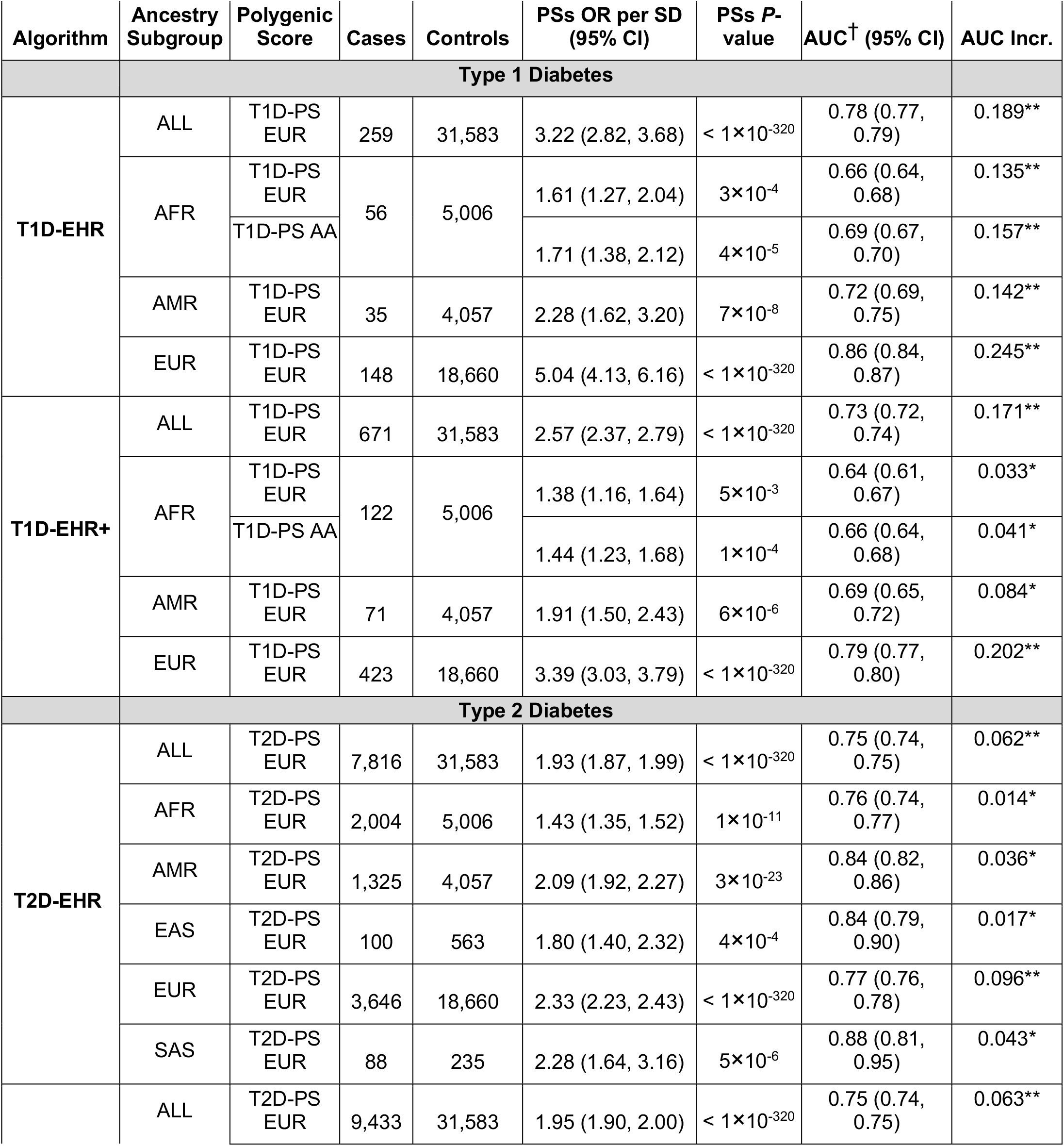
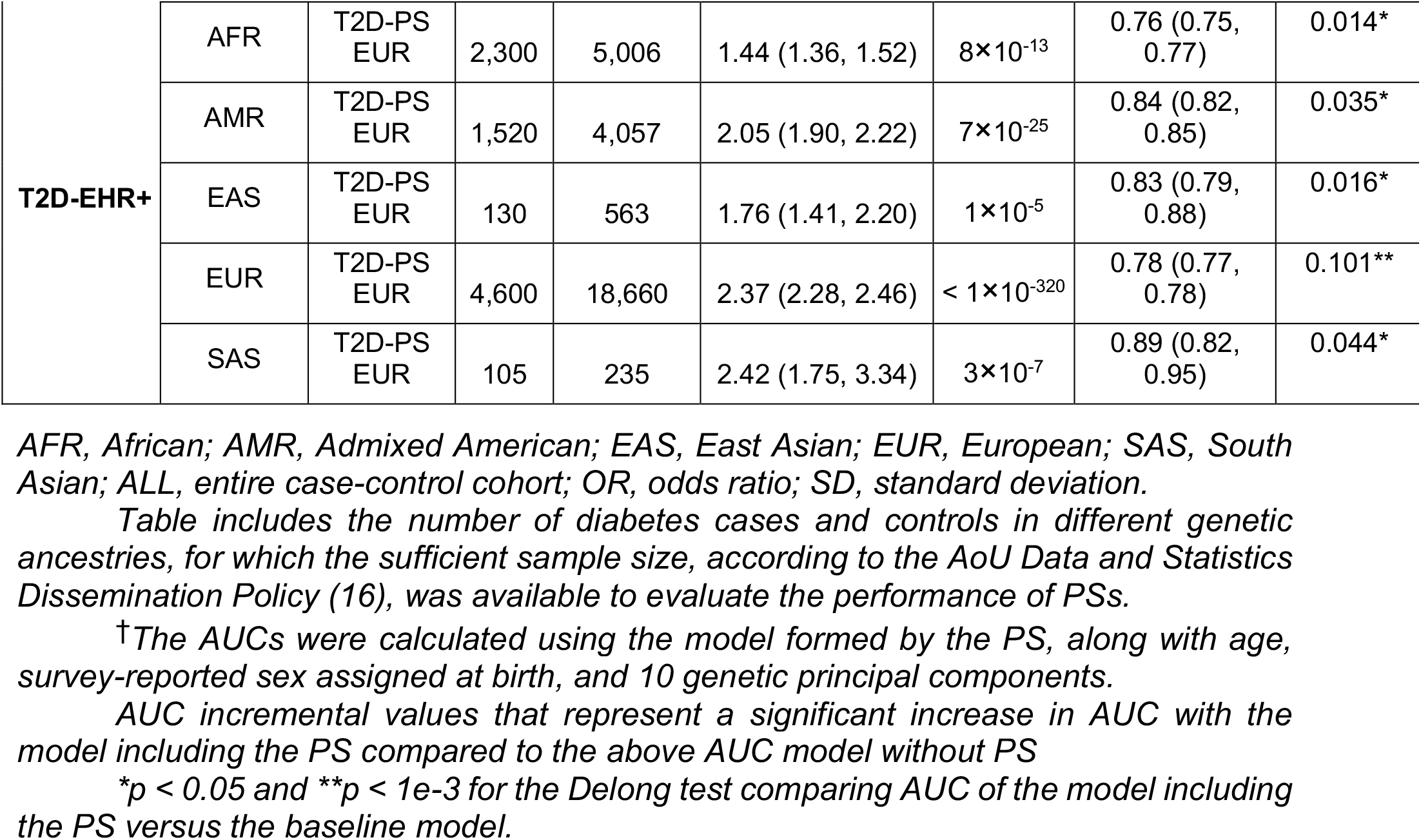
Predictive accuracy of generated PSs using EHR and EHR+ algorithms for cases and universal control algorithm for controls without diabetes identification, in All of Us population, stratified by genetic ancestry subgroups. We exclude genetic ancestry subgroup results for subgroups with insufficient sample size according to the All of Us Data and Statistics Dissemination Policy (16).

**Figure 2.**
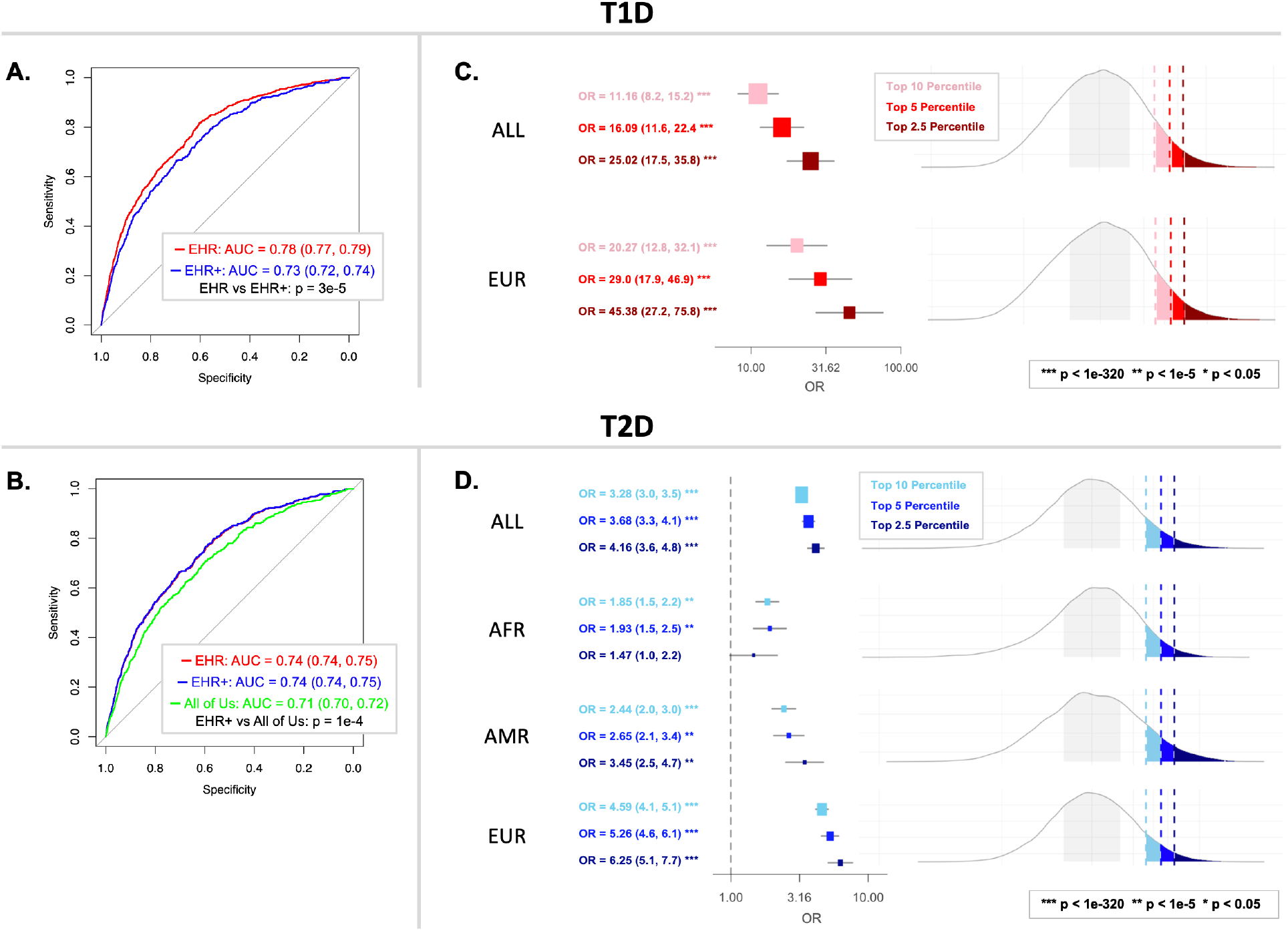
Predictive accuracy of generated PS and diabetes risk discrimination at various percentage cutoffs (2.5%, 5%, or 10%) in AoU population, using developed algorithms. A) Receiver operating characteristic (ROC) curves for T1D-EHR (red) and T1D-EHR+ (blue) definitions, with the p-value of the DeLong test. B) ROC curves for T2D-EHR (red), T2D-EHR+ (blue), and AoU-T2D (green) definitions, with the p-value of the DeLong test, in the entire sample and in genetic-ancestry subgroups with sufficiently large case counts. C) Forest plot for high-risk T1D-PS EUR groups for the T1D-EHR definition (due to the limited number of individuals with genetic data in the AoU cohort (16), we were able to compare the risk of type 1 diabetes defined by developed algorithms only in the European ancestry subgroup). D) Forest plot for high-risk T2D-PS EUR groups for the T2D-EHR+ definition in the entire sample and in genetic-ancestry subgroups with sufficiently large case counts (we did not perform the analysis of extreme T2D-PS EUR thresholds in the East Asian and South Asian ancestry subgroups due to insufficient sample size (16)).

For type 1 diabetes, the T1D-EHR consistently out-performed the T1D-EHR+ algorithm across the full dataset as well as in all analyzed ancestry subgroups, as indicated by larger AUCs (DeLong *p*-value = 3×10^−5^, AUC for entire AoU cohort, **Figure 2A**) and incremental AUC values (**Table 2**), particularly in the European ancestry subgroup. Due to insufficient sample size, we could not evaluate the performance of the T1D-PS EUR in East Asian and South Asian ancestry subgroups.

To evaluate the possible reasons for the superior performance of the T1D-EHR algorithm, we analyzed the clinical characteristics of individuals identified as having type 1 diabetes based on survey data but not EHR data. We observed that across all ancestries, individuals identified as T1D based on survey alone compared to EHR alone had higher BMI (*p*-value = 4.7×10^−26^), were older (*p*-value = 4.8×10^−25^) (**Supplementary Table S4**), which suggest that participants in this group are more likely to have T2D despite self-reporting having T1D.

For type 2 diabetes, the predictive power of the T2D-PS EUR was significantly better when using the T2D-EHR+ algorithm compared to the T2D-EHR or AoU-T2D algorithm in the entire sample (DeLong *p*-values = 0.03 and 1×10^−4^, respectively **Figure 2B, Supplementary Table S5**) and in each of the genetic ancestry subgroups, and was able to identify more cases **(Table 2)**.

We were also interested to see to what extent polygenic scores could classify individuals of diverse ancestries as high risk, using the best-performing type 1 diabetes and type 2 diabetes algorithms, as determined above (T1D-EHR and T2D-EHR+). We looked at risk of diabetes using progressively more extreme cutoffs of the PSs distribution. For type 1 diabetes, in the overall AoU population, individuals within the top 10^th^, 5^th^, 2.5^th^ percentile of the T1D-PS EUR are 11.16, 16.09, and 25.02 times more likely to have disease, compared to the individuals with T1D-PS EUR within the 25^th^-75^th^ percentile (**Figure 2C, Table S6**). For type 2 diabetes, all individuals within the top 10^th^, 5^th^, and 2.5^th^ percentile of the T2D-PS EUR distribution were 3.28, 3.68, and 4.16 times more likely to have type 2 diabetes compared to the individuals with T2D-PS EUR within the 25^th^-75^th^ percentile (**Figure 2D, Table S7**).

## 4. Discussion

To improve the utility of the AoU Research Program in advancing research in diabetes, we have constructed and validated algorithms for diabetes definition, and compared the performance of algorithms that included or excluded participant survey data (EHR vs EHR+). For type 1 diabetes, where no definition was previously available in AoU, we propose that the T1D-EHR algorithm is best performing, identifying more than 250 cases in the AoU cohorts. For type 2 diabetes, the EHR+ definition was best performing, showing increased accuracy and identifying 6,661 (22.57%) more cases than the existing AoU-T2D algorithm.

The generation of accurate phenotype definitions from EHR data is a challenging but crucial step for any type of disease-related research that utilizes large scale biobank data, including the research performed with AoU. Integrating both EHR and self-reported survey data may offer a more complete picture of the individual than either alone, particularly addressing missing information in EHR data. In this study, we use a genetic tool, polygenic scores (12), to validate the accuracy of our newly developed algorithms and to compare across algorithms. Notably, the validity of our diabetes algorithms was confirmed by comparable performance of PSs with established research, both for type 1 (6) and type 2 diabetes (13), supporting the integrity of our case definitions, and underscoring the value of AoU for diabetes research in a diverse population.

While we observed that including survey data increased both the accuracy (as validated by associations with polygenic scores) and the number of cases for type 2 diabetes, we found that for type 1 diabetes, inclusion of survey data resulted in poorer performance. The higher BMI and older age of individuals identified as type 1 diabetes cases based on the survey, but not EHR data, compared to those identified by the EHR algorithm, raises the possibility that some of these individuals could, in fact have type 2 diabetes, suggesting that the inclusion of survey data may inadvertently introduce noise into type 1 diabetes case identification. Thus, for type 1 diabetes, we concluded that the EHR algorithm should be preferentially applied for research in AoU. Whereas for type 2 diabetes, we recommend the preferential use of the EHR+ algorithm, as it substantially increases the number of individuals without introducing contamination of false positives cases.

This study has several strengths, including the development and validation of algorithms that incorporate multiple data sources. We also note that the prediction accuracy of our algorithms showed variations across different populations, highlighting an ongoing Eurocentric bias in genomic studies (14,15). By focusing on diverse groups, the AoU Research Program intends to address this bias.

In conclusion, we provide, for the first time, a validated type 1 diabetes definition for AoU and expand upon existing type 2 diabetes definitions to incorporate both EHR and survey data. We offer access to these harmonized algorithms to help facilitate and standardize diabetes research. Our algorithms, methods, and relevant analytical code will be readily available in the Research Workbench to be shared and implemented by other researchers working within the AoU Research Program.

## Data Availability

All data produced in the present study are available upon reasonable request to the authors

## 5. Acknowledgments

The All of Us Research Program is supported by the National Institutes of Health, Office of the Director: Regional Medical Centers: 1 OT2 OD026549; 1 OT2 OD026554; 1 OT2 OD026557; 1 OT2 OD026556; 1 OT2 OD026550; 1 OT2 OD 026552; 1 OT2 OD026553; 1 OT2 OD026548; 1 OT2 OD026551; 1 OT2 OD026555; IAA #: AOD 16037; Federally Qualified Health Centers: HHSN 263201600085U; Data and Research Center: 5 U2C OD023196; Biobank: 1 U24 OD023121; The Participant Center: U24 OD023176; Participant Technology Systems Center: 1 U24 OD023163; Communications and Engagement: 3 OT2 OD023205; 3 OT2 OD023206; and Community Partners: 1 OT2 OD025277; 3 OT2 OD025315; 1 OT2 OD025337; 1 OT2 OD025276. In addition, the All of Us Research Program would not be possible without the partnership of its participants.

## Funding and Assistance

L.S. is supported by funds from the Ministry of Education and Science of Poland within the project “Excellence Initiative—Research University”, the Ministry of Health of Poland within the project “Center of Artificial Intelligence in Medicine at the Medical University of Bialystok” and American Diabetes Association grant 11-22-PDFPM-03. J.H.L. is supported by NIDDK K23 DK131345 and MGH ECOR Fund for Medical Discovery Clinical Research Award. J.C.F. is supported by NHLBI K24 HL157960. J.M.M. is supported by American Diabetes Association Innovative and Clinical Translational Award 1-19-ICTS-068, American Diabetes Association grant #11-22-ICTSPM-16 and by NHGRI U01HG011723. A.K.M. is supported by the Foundation for the National Institutes of Health with funding from AMP CMD RFP 2: GENERATION of New genetic, -omic, or biomarker data for Common Metabolic Diseases titled “Common metabolic disease genetic association analysis in the All of Us Research Program” and by NHGRI U01HG011723. M.S.U. is supported by NIDDK K23DK114551, NIDDK R03DK131249, and Doris Duke Foundation Award 2022063.

## Author Contributions and Guarantor Statement

L.S., R.M. and P.S. researched data, wrote, reviewed, and edited the manuscript. B.C.P., J. H. L. and J. C. F. reviewed and edited the manuscript. J. M. M., A.K.M. and M.S.U. reviewed and edited the manuscript and are the guarantors of this work and, as such, had full access to all the data in the study and take responsibility for the integrity of the data and the accuracy of the data analysis.

## Prior Presentation

Part of the results included in this article was presented at the All of Us Researchers Convention (March 30^th^, 2023).

