## Supplemental material for "Algorithms for the identification of prevalent diabetes in the All of Us Research Program validated using polygenic scores – a new resource for diabetes precision medicine"

### Supplementary methods:

#### A. Cohort description and data extraction

A detailed description of the AoU Research Program, including goals, recruitment methods, clinical sites, data availability and storage, has been previously published (1). We analyzed data from 372,397 AoU participants enrolled by January 1, 2022. Among them, 352,366 consented to submit EHR data, and 309,974 people already had EHR data available (in the form of a reported condition, device implantation, drug exposure, measurement, procedure, or visit). On June 22, 2022, the AoU Research Program released whole genome sequencing (WGS) data for 98,590 study participants. EHR, survey, and genetic information was extracted from the AoU v6 controlled tier dataset on November 3, 2022.

The AoU Research Program provides a list of individuals flagged as having low quality WGS data based on number of variants, insertion/deletion ratio, number of insertions, number of deletions, variant and indel heterozygosity, transition/transversion ratio, and the number of variants not in gnomAD v3.1. We removed these flagged individuals from our genetic analyses.

Genetic ancestry classifications were defined by the AoU Research Program. Briefly, a random forest classifier predicting ancestry classifications from principal components (PCs) using chr20 and chr21 variants from the Human Genome Diversity Project (HGDP) and 1000 Genomes (1000G) was created. Then, a set of linkage disequilibrium (LD)-pruned with a $r^2$ cutoff of 0.1, bi-allelic variants with allele frequency >0.1% and call rate >0.99% from the AoU WGS data were used to project PCs into the same HGDP/1000G PC space. The classifier was then applied to the AoU data to obtain ancestry classification probabilities, and hard classifications were determined as the continental ancestry with a probability over 75%. The resulting subgroups were labeled as African/African American (AFR), American Admixed/Latino (AMR), East Asian (EAS); European (EUR) and South Asian (SAS).

For the algorithms, described below, we incorporated multiple data points available in AoU. An EHR recorded diagnosis was defined as the presence of an ICD9 or ICD10 code of diabetes (diagnosed by a clinician) in the EHR, and we also considered self-reported diabetes diagnosis (survey question: “Has a doctor or health care provider ever told you that you have Type 1 Diabetes / Type 2 Diabetes / Other / UNKN diabetes?”). Diabetes medications were analyzed using medication names that were translated to universal codes using the anatomical therapeutic chemical (ATC) classification (2). Laboratory data was extracted using the Logical Observation Identifiers Names and Codes (LOINC) – a standardized method to code laboratory test orders and results (3). Self-reported race, date of birth, and gender were pulled from the AoU survey data, and age was calculated as of November 3, 2022. LOINC codes and QC steps used to clean the BMI, HbA1c, random glucose, and fasting glucose data is in **Supplementary Table S1**.

### B. Development of diabetes phenotyping algorithms

Our goal was to develop algorithms to identify individuals diagnosed with type 1 diabetes or type 2 diabetes, as well as individuals without a diabetes diagnosis. These sets of individuals could serve as "cases" and "controls" for diabetes studies. We created two versions of case identification algorithms: the EHR and EHR+ algorithms. The EHR algorithms utilized EHR-driven diagnosis, medications, and laboratory measurement values and were developed from previously reported algorithms (4,5). The EHR+ algorithm also included self-reported diagnosis provided by survey data. We used the American Diabetes Association's (ADA) diagnostic criteria for diabetes (6): fasting glucose ≥ 126 mg/dl and/or HbA1c ≥ 6.5% (48 mmol/mol). ADA criteria also includes the random glucose criterion, where diabetes can be diagnosed when the concentration is ≥ 200 mg/dl, but this requires the presence of classic symptoms of hyperglycemia or hyperglycemic crisis, which is not readily available in EHR data. Therefore, we did not use random glucose in the type 2 diabetes case selection algorithm. We kept the exclusion criterion of random glucose concentration ≥ 200 mg/dl in the selection of individuals without a diabetes diagnosis, as the inclusion of individuals with abnormal random glucose records could potentially introduce individuals with undiagnosed diabetes.

For the type 1 diabetes case identification algorithm, we modified the algorithm proposed by the eMERGE Phase-IV Program (4). Briefly, patients with an ICD code for type 1 diabetes and who had been prescribed insulin in an outpatient setting were identified. Note that we ensured that medications were prescribed in the outpatient vs inpatient setting given the high rate of insulin use in hospitalized patients with all forms of diabetes. Patients were then excluded if they had any non-insulin diabetes medication prescriptions, or had a diagnosis of malignant cancer, cystic fibrosis, or drug-induced diabetes, as defined by ICD codes. We labeled this algorithm 'T1D-EHR.' An additional type 1 diabetes EHR+ (labeled 'T1D-EHR+') definition was created, including individuals designated as a case by the EHR algorithm described above or who responded as having type 1 diabetes in their participant survey. These algorithms for type 1 diabetes case identification are presented in **Figure 1A**.

For the type 2 diabetes case identification algorithm, we modified the Northwestern University algorithm (5). In the first step, we filtered out individuals with an ICD-based diagnosis of type 1 diabetes. After this exclusion step, there are multiple possible decision paths to code a participant as having type 2 diabetes. First, we required cases with a type 2 diabetes diagnosis based on ICD codes to have a prescription of diabetes medications. If a patient has an outpatient insulin treatment record, either the prescription of at least one additional non-insulin diabetes medication before initiation of insulin therapy or at least two different independent ICD diagnoses of type 2 diabetes are required for the individual to be classified as a type 2 diabetes case. For individuals with type 2 diabetes ICD-based diagnosis but without any diabetes medication records, we required the presence of at least one abnormal glycemic laboratory result (fasting glucose ≥ 126 mg/dl or HbA1c ≥ 6.5%) before assigning them as a type 2 diabetes case. Finally, for individuals without a type 2 diabetes diagnosis codes in the EHR, we required a prescription for any non-insulin diabetes medication and abnormal laboratory results for fasting glucose or HbA1c before assigning them as a type 2 diabetes case. We labeled this algorithm 'T2D-EHR.' A second type 2 diabetes case definition, EHR+ (labeled 'T2D-EHR+') was created, which additionally included individuals who responded as having type 2 diabetes in their participant survey. The algorithm for type 2 cases identification is presented in **Figure 1B**. Finally, we applied the algorithm for type 2 diabetes case identification available in the "Phenotype Library" of the AoU Researcher Workbench "Featured Workspaces" (labeled 'AoU-T2D'). This algorithm is similar to ours, based on the Northwestern University algorithm, but most notably differs from our implementation by 1) the usage of random glucose as an abnormal glycemic lab to assign type 2 diabetes case status, 2) the use of diabetes medications without filtering for visit settings (in- or outpatient), and 3) not classifying participants with the following conditions (a T2D ICD code AND no T2D or T1D medications AND no T1D ICD codes AND with abnormal labs) as a case. Details of the AoU-T2D case algorithm are presented below in the section "Description of the existing in AoU Phenotyping Library T2D algorithm".

We defined a universal algorithm for the identification of control individuals without diabetes (**Figure 1C**), based on the Northwestern University type 2 diabetes control algorithm (16). Participants were included as controls if they had 1) at least one in-person healthcare provider visit, 2) at least one glucose measurement available in the EHR, 3) no abnormal random glucose, fasting glucose, or HbA1c lab values if they are available, 4) no diagnoses of any type of diabetes or related conditions (prediabetes, glycosuria), based on EHR data or survey responses, and 5) no prescriptions of diabetes medications. This differs from the published Northwestern University type 2 diabetes control algorithm which requires at least two in-person healthcare visits, removes participants with a family history of diabetes, does not include survey information in their inclusion criteria, and defines abnormal random glucose, fasting glucose, and HbA1c lab values as ≥110 mg/dl, ≥110 mg/dl, and ≥6.0% respectively.

### C. Polygenic scores for diabetes

We implemented two published type 1 diabetes polygenic scores. First, we applied the polygenic score model published by Sharp *et al.* (7), which consisted of 67 single nucleotide polymorphisms (SNPs) (including 14 HLA DR-DQ, 21 other HLA, and 32 non-HLA SNPs), using the steps outlined in the HLA PRS Toolkit (8). This PS, labeled 'T1D-PS EUR', was derived from genetic associations in predominantly European ancestry cohorts. Second, we applied a type 1 diabetes score comprised of 12 variants with weights derived from genetic associations in African-American cohorts published by Onengut-Gumuscu *et al*. (9). We labeled this score 'T1D-PS AA'.

We generated a global extended type 2 diabetes polygenic score using the PRS-CS software (10) with the auto option used to set the global shrinkage parameter. As input to the software, we meta-analyzed summary statistics from the European ancestry subset GWAS meta-analysis of the type 2 diabetes published by Vujkovic *et al.* (11) and the type 2 diabetes genome-wide association study made publicly available by the FINNGEN Consortium (12) (Release 6). The summary statistics had a final sample size of 277,802 type 2 diabetes cases and 1,434,249 controls. The meta-analysis was performed using METAL (13), with inverse-variance weighting. For PRS-CS, we used the linkage disequilibrium reference panel from the UK Biobank, available from the PRS-CS github page (14). We labeled this score the 'T2D-PS EUR'.

For all polygenic scores calculated, we applied the ancestry adjustment methods described in Khera *et al.* (15) using the genetic ancestry probabilities extracted from AoU.

This adjustment involved fitting a linear regression model using the inferred probabilities of being assigned to each ancestry category with the raw polygenic score as the response variable. The residuals from this model were the 'ancestry-adjusted' values used in all subsequent analyses.

To assess the accuracy of our diabetes definitions in the context of polygenic scores, we performed a logistic regression analysis with case-control status as the outcome and the disease-specific polygenic score as the dependent variable. We constructed regression models that included age, sex, and genetic PCs as covariates with and without inclusion of the PS. The models with the ancestry-adjusted PSs did not include genetic PCs, as the PSs were already ancestry-adjusted. For each diabetes definition, we evaluated the area under the receiver operating characteristic curve (AUC) of the full model including the PS. We also evaluated the incremental AUC, calculated as the decrease in AUC when removing the PS from the model. We evaluated the significance of the difference between the AUC of the model with and without the PS using the DeLong test (16). To assess the effect of the PS at the extreme tail of the PSs, we identified individuals at the top 10%, 5%, or 2.5% of the PSs distribution, and calculated the odds ratio (OR) per standard deviation of PSs of these high-risk individuals compared to individuals within the $25^{th}$-$75^{th}$ percentile of the samples, adjusting for the covariates.

### D. Description of the existing in AoU Phenotyping Library type 2 diabetes algorithm:

1. 1 or more ICD codes for type 2 diabetes AND 1 or more type 2 diabetes medications excluding individuals with 1 or more T1D ICD code.
    1. type 2 diabetes medications from search of any medication containing the following words:
    1. 'acetohexamide', 'tolazamide', 'chlorpropamide', 'glipizide', 'glyburide', 'glimepiride','repaglinide', 'nateglinide', 'metformin', 'rosiglitazone', 'pioglitazone', 'troglitazone', 'acarbose', 'miglitol', 'sitagliptin', 'exenatide', 'saxagliptin', 'linagliptin', 'liraglutide', 'semaglutide', 'canagliflozin', 'dapagliflozin', 'empagliflozin', 'alogliptin', 'colesevelam', 'albiglutide', 'dulaglutide', 'lixisenatide'
    2. No further filtering for mixed medications
2. 1 or more type 2 diabetes ICD code and 2 or more type 1 and type 2 diabetes medications with type 2 diabetes medication coming >= 1 day before type 1 diabetes medication, excluding individuals with 1 or more type 1 diabetes ICD code.
1. type 1 diabetes medications from search of any medication containing the following words:
    1. 'insulin', 'pramlintide', 'insulin glulisine', 'insulin lispro', 'insulin aspart', 'insulin glargine', 'insulin detemir', 'insulin degludec', 'insulin NPH'
    2. No filtering of in-patient vs out-patient
    3. No further filtering for mixed medications
3. 2 or more type 2 diabetes ICD codes and 1 or more type 1 diabetes medication, excluding individuals with 1 or more T1D ICD code.
4. 1 or more type 2 diabetes medication and 1 or more abnormal lab, excluding individuals with 1 or more type 1 diabetes ICD code.
1. Abnormal labs include:
    1. Random Glucose > 200 mg/dl
    2. Fasting glucose >= 125 mg/dl
    3. A1c >= 6.5%
    4. No filtering of unusual units

### Supplementary tables:

**Supplementary Table S1.** LOINC codes and QC steps used to clean the BMI, HbA1c, random glucose, and fasting glucose data.

| Name | LOINC codes | Units selected | Additional filtering |
|---|---|---|---|
| BMI | 39156-5 | kilogram per square meter, kg/sq. m | remove values >2sd away from average; remove values >=100; take average of measurements from the last 5 years |
| HbA1C | 4548-4, 17856-6,4549-2, 17855-8 | percent, Percent, Of Total H, Percentage unit, percent hemoglobin, percent hemoglobin A1c, % of total | remove values >=20 |
| Random Glucose | 2339-0, 2345-7 | milligram per deciliter, mg/dL | |
| Fasting Glucose | 1558-6 | milligram per deciliter, mg/dL | |

**Supplementary Table S2.** The demographics of identified type 2 diabetes cases using the existing in AoU Phenotyping Library type 2 diabetes algorithm, stratified by self-reported race.

| | *Type 2 diabetes AoU Phenotyping Library algorithm* | | | | | |
|---|---|---|---|---|---|---|
| | **All** | **NHW** | **NHB** | **HIS** | **ASIAN** | **Other** |
| **N** | 29511 | 12827 | 7799 | 6444 | 542 | 1899 |
| **Age, mean (SD)** | 63.61 (12.65) | 67.12 (12.34) | 61.06 (11.31) | 59.81 (12.68) | 62.83 (14.08) | 63.44 (13.04) |
| **BMI kg/m$^2$, mean (SD)** | 33.96 (8.34) | 34.0 (8.02) | 35.11 (9.23) | 33.01 (7.6) | 28.35 (6.18) | 33.77 (8.31) |
| **N, Female (%)** | 16744 (56.74%) | 6554 (51.1%) | 5086 (65.21%) | 4008 (62.2%) | 284 (52.4%) | 812 (42.76%) |
| **HbA1c %, mean (SD) [mmol/mol, mean (SD)]** | 7.32 (1.95) [56.47 (21.33)] | 7.43 (2.04) [57.66 (22.31)] | 7.04 (1.66) [53.4 (18.12)] | 7.21 (1.87) [55.28 (20.44)] | 7.51 (1.94) [58.53 (21.25)] | 7.24 (1.93) [55.62 (21.15)] |

**Supplementary Table S3.** The demographics of identified control without diabetes subjects, using the developed universal control algorithm, stratified by self-reported race.

| | ***Universal control algorithm*** | | | | | |
|---|---|---|---|---|---|---|
| | **All** | **NHW** | **NHB** | **HIS** | **ASIAN** | **Other** |
| **N** | 89563 | 54203 | 12757 | 14687 | 2066 | 5850 |
| **Age, mean (SD)** | 53.82 (17.31) | 57.79 (17.21) | 50.81 (14.93) | 43.68 (14.42) | 45.35 (14.82) | 52.12 (17.54) |
| **BMI kg/m$^2$, mean (SD)** | 28.6 (6.97) | 28.06 (6.57) | 30.06 (8.17) | 29.97 (7.03) | 24.48 (4.57) | 28.39 (7.04) |
| **N, Female (%)** | 56675 (63.28%) | 34506 (63.66%) | 7593 (59.52%) | 10519 (71.62%) | 1363 (65.97%) | 2694 (46.05%) |
| **HbA1c %, mean (SD) [mmol/mol, mean (SD)]** | 5.21 (0.29) [33.46 (3.13)] | 5.21 (0.3) [33.49 (3.23)] | 5.22 (0.27) [33.59 (2.97)] | 5.2 (0.28) [33.28 (3.05)] | 5.2 (0.28) [33.34 (3.07)] | 5.21 (0.28) [33.45 (3.09)] |

**Supplementary Table S4.** The demographics of individuals identified as type 1 diabetes cases by the survey response, but not the T1D-EHR algorithm, stratified by self-reported race.

| | **All** | **NHW** | **NHB** | **HIS** | **ASIAN** | **Other** |
|---|---|---|---|---|---|---|
| **N** | 1618 | 887 | 269 | 287 | 49 | 126 |
| **Age, mean (SD)** | 56.83 (15.78) | 58.88 (15.48) | 54.0 (15.62) | 53.85 (16.14) | 56.47 (14.31) | 55.44 (16.0) |
| **BMI kg/m$^2$, mean (SD)** | 31.69 (8.48) | 31.8 (8.38) | 32.0 (8.71) | 31.75 (9.0) | 29.76 (7.0) | 31.0 (8.05) |
| **N, Female (%)** | 912 (56.37%) | 307 (34.61%) | 197 (73.23%) | 281 (97.91%) | 48 (97.96%) | 79 (62.7%) |
| **HbA1c %, mean (SD) [mmol/mol, mean (SD)]** | 7.5 (1.92) [58.44 (21.03)] | 7.38 (1.9) [57.12 (20.79)] | 7.75 (2.14) [61.22 (23.41)] | 7.72 (1.98) [60.88 (21.66)] | 6.95 (1.3) [52.46 (14.21)] | 7.49 (1.55) [58.38 (16.97)] |
| **Number (%) of individuals diagnoses as T2D based on T2D-EHR algorithm** | 327 (20.21) | 191 (21.53) | 44 (16.36) | 60 (20.91) | 14 (28.57) | 18 (14.29) |

**Supplementary Table S5**. Predictive accuracy of generated T2D-PS using the existing in AoU Phenotyping Library type 2 diabetes algorithm, in AoU population, stratified by genetic ancestry.

| Type 2 diabetes Definition | Genetic Ancestry | Cases | Controls | OR (95% CI) | P | AUC (95% CI) |
|---|---|---|---|---|---|---|
| ***Type 2 diabetes AoU Phenotyping Library algorithm*** | **ALL** | 7,948 | 31,583 | 1.91 (1.85, 1.97) | $< 1*10^{-320}$ | 0.71 (0.70, 0.72) |
| | **AFR** | 2,081 | 5,006 | 1.39 (1.30, 1.48) | $5*10^{-11}$ | 0.72 (0.71, 0.73) |
| | **AMR** | 1,318 | 4,057 | 2.0 (1.85, 2.15) | $3*10^{-19}$ | 0.79 (0.77, 0.81) |
| | **EAS** | 95 | 563 | 1.70 (1.33, 2.06) | $7*10^{-3}$ | 0.78 (0.73, 0.83) |
| | **EUR** | 3,720 | 18,660 | 2.32 (2.22, 2.42) | $< 1*10^{-320}$ | 0.73 (0.72, 0.74) |
| | **SAS** | 87 | 235 | 2.34 (1.66, 3.27) | $8*10^{-5}$ | 0.84 (0.78, 0.90) |

**Supplementary Table S6.** Number of identified cases in individuals at the top 2.5%, 5%, or 10% of the PSs distribution for type 1 diabetes, using EHR and EHR+ algorithms.

| Used Algorithm | Genetic Ancestry | Percentile of PS | Number of Cases | Number of Controls |
|---|---|---|---|---|
| **T1D-EHR** | **ALL** | top 10 | 127 | 3,008 |
| | | top 5 | 92 | 1,516 |
| | | top 2.5 | 67 | 715 |
| | **EUR** | top 10 | 92 | 1,802 |
| | | top 5 | 66 | 905 |
| | | top 2.5 | 46 | 410 |
| **T1D-EHR+** | **ALL** | top 10 | 280 | 3,008 |
| | | top 5 | 193 | 1,516 |
| | | top 2.5 | 134 | 715 |
| | **EUR** | top 10 | 212 | 1,802 |
| | | top 5 | 146 | 905 |
| | | top 2.5 | 98 | 410 |

**Supplementary Table S7.** Numbers of identified cases in individuals at the top 2.5%, 5%, or 10% of the PSs distribution for type 2 diabetes, using EHR and EHR+ algorithms.

| Used Algorithm | Genetic Ancestry | Percentile of PS | Number of Cases | Number of Controls |
|---|---|---|---|---|
| T2D-EHR | ALL | top 10 | 1,486 | 2,351 |
| | | top 5 | 763 | 1,082 |
| | | top 2.5 | 411 | 528 |
| | AFR | top 10 | 255 | 377 |
| | | top 5 | 118 | 148 |
| | | top 2.5 | 49 | 76 |
| | AMR | top 10 | 286 | 466 |
| | | top 5 | 161 | 252 |
| | | top 2.5 | 105 | 134 |
| | EUR | top 10 | 780 | 1,200 |
| | | top 5 | 385 | 525 |
| | | top 2.5 | 191 | 226 |
| T2D-EHR+ | ALL | top 10 | 1,806 | 2,351 |
| | | top 5 | 936 | 1,082 |
| | | top 2.5 | 500 | 528 |
| | AFR | top 10 | 285 | 377 |
| | | top 5 | 128 | 148 |
| | | top 2.5 | 52 | 76 |
| | AMR | top 10 | 318 | 466 |
| | | top 5 | 183 | 252 |
| | | top 2.5 | 117 | 134 |
| | EUR | top 10 | 1007 | 1,200 |
| | | top 5 | 507 | 525 |
| | | top 2.5 | 255 | 226 |
